# In-vivo glioma viscosity and fluidity as clinical tumor markers of vimentin expression and collective cell migration

**DOI:** 10.64898/2026.06.21.26356180

**Authors:** Mehrgan Shahryari, Pablo Gottheil, Helge Herthum, Tom Meyer, Elisabeth G. Hain, Jörg Schnauß, Eberhard Siebert, Vincent Prinz, Josef Käs, Ingolf Sack

## Abstract

Reduced fluidity and viscosity have been demonstrated as biomechanical hallmarks of in vivo glioblastoma and are increasingly used as radiological imaging markers by magnetic resonance elastography (MRE). However, the biological origin and consequences of this unusual mechanical behavior remain unclear. Here, we show that two mechanisms which promote collective cell migration are present in patient gliomas and can be detected in vivo by MRE-based cerebral tomoelastography. Vimentin-driven extracellular matrix remodeling and cellular elongation, quantified by automated histological readings and nuclear aspect ratio (AR) measurements, correlate with decreased in-vivo tumor fluidity and viscosity. These observations in patients are supported by experiments in tissue-mimicking actin–vimentin gels, which mechanistically link the soft-solid viscoelastic signature of in vivo glioma to vimentin’s migration-promoting role and to AR-based observations of cellular elongation in unjammed cancer cell clusters. Taken together, our results suggest in-vivo bulk tumor viscosity as a noninvasive biomechanical marker of collective cell migration and invasiveness in brain tumors.

## Introduction

Glioma is one of the most devastating and fatal tumors, with a median overall survival of 14-15 months for high-grade gliomas.(*1*) Despite recent advances in anti-tumor therapies, glioma treatment suffers from insufficient diagnostic tools.(*2*) Although non-invasive imaging markers can detect and differentiate malignant masses in the brain, histology-based tissue assessment remains the gold standard, while patient-specific predictions of treatment resistance or recurrence after therapy often fail.(*2, 3*) In addition, imaging-based detection of tumor margins before surgery and pseudoprogression after radiotherapy remain major hurdles in glioma diagnostics.(*4*)

Understanding the physical properties of gliomas may be key to improving therapeutic outcomes as well as the specificity and predictivity of diagnostic imaging.(*5, 6*) For example, tumor biomechanical properties have been shown to play a critical role in shaping the tumor niche, promoting tumor progression, facilitating metastasis, and fostering therapeutic resistance.(*7–13*)

However, most of these reports are based on biomechanical assays and testing methods that are not applicable to patients. In vivo mechanical properties of the brain can currently only be obtained non-invasively and with high spatial resolution using magnetic resonance elastography (MRE).(*14*) MRE encodes externally induced shear waves by three-dimensional phase-contrast MRI as a probe of soft tissue viscoelastic properties.(*15*) Although cerebral MRE has been developed for more than a decade, there remain challenges in high-resolution mapping of sufficient consistency. In particular, in brain tumors, MRE has not yet fully exploited the rich information conveyed by biomechanical parameters.(*16*) Current MRE reports on gliomas agree on the surprisingly high heterogeneity in stiffness (often including stiff solid and fluid viscous regions in the same lesion),(*17*) softer properties than normal-appearing brain matter (covering a wide range of stiffness values),(*18–22*) and low values of relative viscosity(*23*) also referred to as tissue fluidity (resulting in solid yet very soft tissue properties).(*5*) To date, a consistent multiparametric MRE assessment of glioma based on stiffness, tissue fluidity, and absolute viscosity using multifrequency MRE and histopathologic quantification is still pending.

At the microscopic level, tumor aggressiveness is associated with the motility and invasiveness of cancer cells. From a physics perspective, cancer cells must generate the forces that allow them to unjam, pull on their stroma, and change position.(*11, 24–26*) These processes, which are synonymous with micro-tissue fluidization, require that cells act cooperatively, e.g., by forming multicellular streams through the remodeled extracellular matrix (ECM) even though the tumor as a whole may behave as a stiff-rigid entity.(*13*) Strikingly, the motility of cancer cells has been associated with characteristic cell and nucleus shape changes, such as the nucleus aspect ratio (AR), as a marker for tumor aggressiveness.(*27*) However, it remains unclear whether cancer cell motility and the tumor microenvironment at the tissue level give rise to a distinct biomechanical profile that can be exploited as an MRE-based biomarker in glioma.

To address this question, we first identified the intermediate filament vimentin as a key regulator of cytoskeletal force transmission and traction force generation promoting cancer cell motility.(*28, 29*) Through adjustments in solubility, polymer chain length, and cross-linking, vimentin possesses a powerful machinery to dynamically adapt the cytoskeleton to large strains while maintaining soft-elastic cell properties in response to substrate stiffness.(*30, 31*) This special biomechanical function is associated with unusual viscoelastic properties not shared by other filamentous biopolymers such as actin or tubulin, which rupture at relatively low levels of strain.(*32, 33*) In metastatic cancer, vimentin has been identified as a protector against oxidative stress(*34*) and is used as a histopathological marker of epithelial-mesenchymal transformation indicating poor prognosis.(*35*) The mechano-biological function of vimentin in promoting cancer cell invasiveness combined with its unique biomechanical fingerprint may make MRE a highly desirable non-invasive imaging marker for glioma detection and staging.

Therefore, we here combine multifrequency-MRE based tomoelastography(*36*) with quantitative histology and cell nucleus shape analysis in a group of glioma patients. Tomoelastography is an emerging imaging technique that yields highly spatially consistent, multiparametric biomechanical maps of shear wave speed (SWS, m/s), penetration rate (PR, m/s), and the phase angle of the complex shear modulus (φ, rad). We considered these in vivo parameters as proxies for glioma stiffness, inverse absolute viscosity, and tissue fluidity, respectively. Since PR and φ capture absolute and relative tissue viscosity, they are discussed jointly as MRE-viscosity.(*37, 38*) Our goal was to identify which of these properties is sensitive to the biomechanical signature of cancer cell migration in glioma. Ultimately, as a non-invasive clinical technique, tomoelastography can harness emergent biomechanical signatures of cancer cells and their microenvironment to bridge the gap between macroscopic imaging and micromechanical changes underlying collective cell migration.

## Results

### Vimentin decreases tissue fluidity in phantoms

Networks with varying actin and vimentin compositions demonstrated significant frequency-dependent changes of SWS, PR and *φ* across the 1 to 10 Hz frequency range (all p < 0.001; Figure 1a). SWS, PR, and *φ* differed significantly among the five networks with varying actin and vimentin compositions when frequency data were pooled (all p < 0.001), reflecting distinct mechanical properties across the different vimentin compositions (Figure 1b). Notably, from 0 to 1 g/L vimentin phantom concentration, PR increased with approximately 260% (0.028 ± 0.002 m/s to 0.102 ± 0.008 m/s) while *φ* decreased by 67% (0.512 ± 0.072 rad to 0.169 ± 0.019 rad). SWS increased only by 16% (0.046 ± 0.004 m/s to 0.054 ± 0.002 m/s).

**Figure 1.**
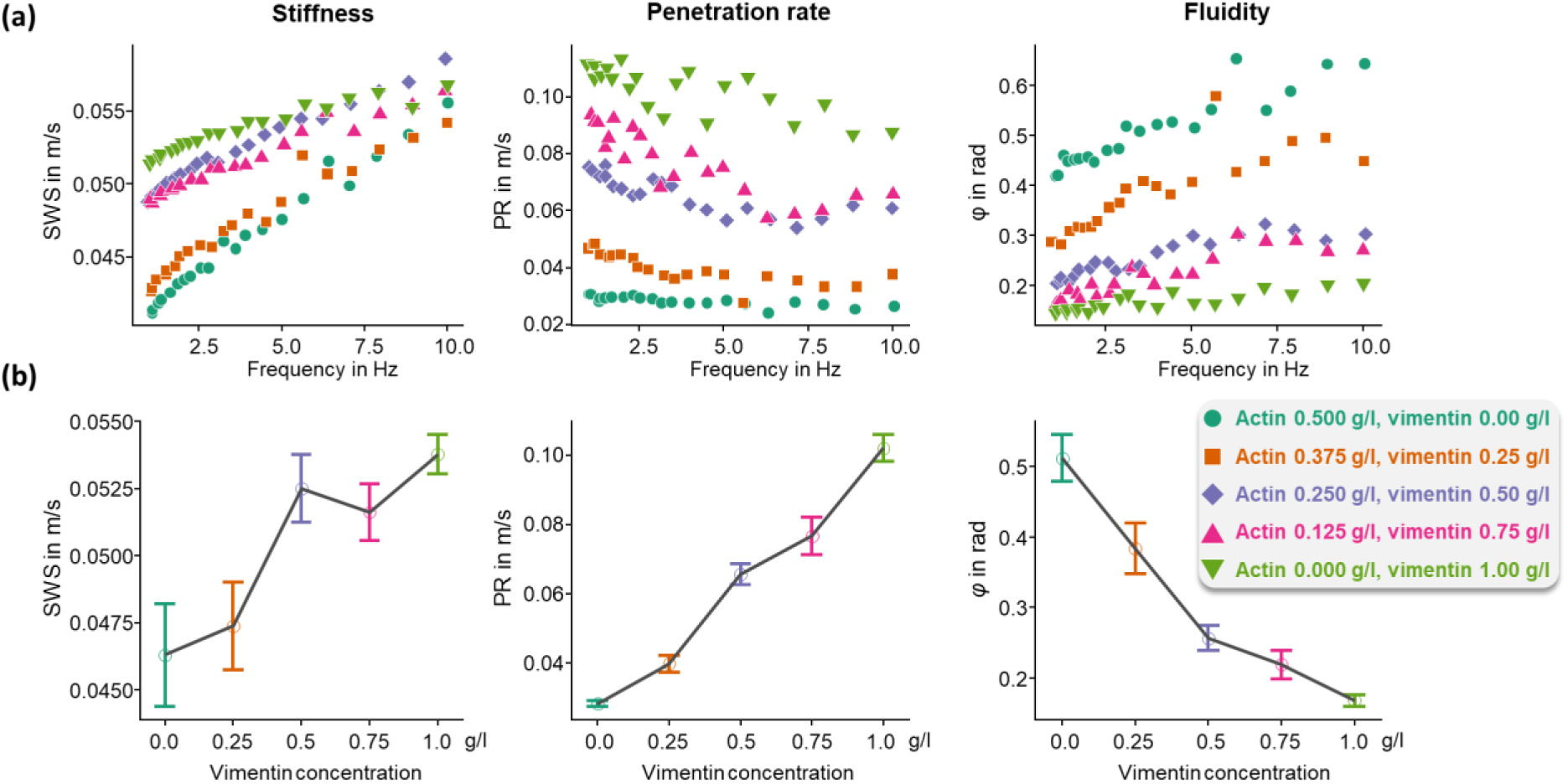
Mechanical properties of in vitro networks with varying actin and vimentin compositions based on shear rheometry. **(a)** Frequency-dependent relationships (1–10 Hz) for shear wave speed (SWS), penetration rate (PR), and loss angle (*φ*) across the five network compositions with vimentin concentrations ranging from 0 to 1 g/L. Symbols represent mean values; all parameters showed significant frequency dependence (all p < 0.001). **(b)** Frequency-averaged mean values of SWS, PR, and *φ* for the five network compositions with vimentin concentrations 0 to 1 g/L. Kruskal–Wallis testing revealed significant differences among compositions for the three mechanical parameters used in our MRE study (all p<0.001).

### Tomoelastography reveals spatial details in glioma stiffness and viscosity markers

Cerebral tomoelastography was successfully performed in all patients without technical failure, enabling tumor delineation in all cases. Patient characteristics and mean values of SWS, PR, *φ*, and ADC, as well as quantified histological analysis of glioma samples, are summarized in Table 1. Figure 2 shows representative vimentin staining from three glioma cases - a grade 2 astrocytoma with low vimentin expression, a glioblastoma with moderate vimentin expression, and a glioblastoma with high vimentin expression - alongside the corresponding MRI and tomoelastography maps. Zoomed inserts highlight the unprecedented resolution of biomechanical detail revealed by tomoelastography.

**Figure 2:**
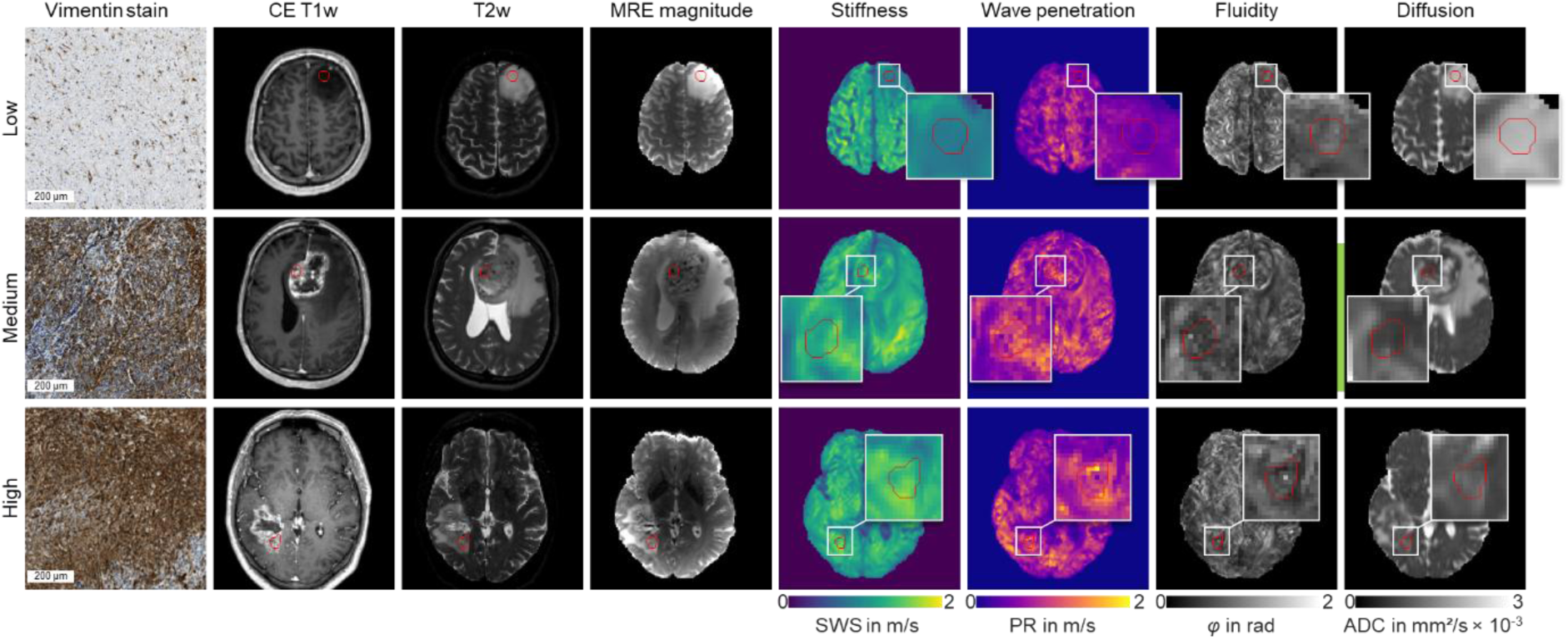
Vimentin expression, anatomical MRI and tomoelastography maps, and DWI in gliomas. Representative vimentin immunostaining (620 × 620 µm) from three gliomas: a grade 2 astrocytoma with low vimentin expression, a glioblastoma with moderate vimentin expression, and a glioblastoma with high vimentin expression. Mean values ± standard deviation for low, moderate, and high vimentin expression were: Shear wave speed (SWS), 0.99 ± 0.06 m/s, 1.30 ± 0.12 m/s, and 1.43 ± 0.14 m/s, respectively; penetration rate (PR), 0.70 ± 0.19 m/s, 0.99 ± 0.23 m/s, and 1.09 ± 0.30 m/s, respectively; loss angle (*φ*), 0.47 ± 0.11 rad, 0.43 ± 0.10 rad, and 0.44 ± 0.12 rad, respectively; and apparent diffusion coefficient (ADC), 913 ± 114, 831 ± 125, and 1069 ± 102 × 10⁻⁶ mm²/s, respectively.

**Table 1.** Patient description with key imaging and histological parameters. Means are given with ± SD, range or IQR as shown. Shear wave speed (SWS), penetration rate (PR), loss angle (*φ*), apparent diffusion coefficient (ADC), nuclear aspect ratio (AR).

| Group | Low | High |
| --- | --- | --- |
| WHO-Grade | 2 and 3 | 4 |
| Participant numbers | 6 | 17 |
| Entity | 2 astrocytoma and 2 oligodendroglioma (grade 2);<br>1 astrocytoma and 1 oligodendroglioma (grade 3) | 1 IDH-mutant astrocytoma (grade 4);<br>13 glioblastoma and 3 molecular glioblastoma (grade 4) |
| Sex female | 3 | 5 |
| Age in years (range) | 42 (24-61) | 55 (33-67) |
| BMI in kg/m<sup>2</sup> (IQR) | 26.2 ± 3.7 (25.6 - 28.6) | 28.3 ± 6.4 (23.1-30.4) |
| SWS in m/s (IQR) | 1.17 ± 0.25 (1.06 - 1.28) | 1.16 ± 0.12 (1.09 - 1.23) |
| PR in m/s (IQR) | 0.7 ± 0.21 (0.55 - 0.86) | 0.81 ± 0.15 (0.75 - 0.88) |
| $\varphi$ in rad (IQR) | 0.56 ± 0.1 (0.5 -0.59) | 0.48 ± 0.07 (0.44 - 0.52) |
| ADC in mm<sup>2</sup>/s*10<sup>-6</sup> (IQR) | 1521 ± 338 (1332 - 1570) | 1073 ± 245 (913 - 1173) |
| Mean nuclear AR (IQR) | 1.43 ± 0.07 (1.39 - 1.44) | 1.53 ± 0.09 (1.45 - 1.59) |
| Reticulin signal per 1/mm<sup>2</sup> (IQR) | 1.48e-05 ± 1.00e-05 (1.04e-05 - 1.96e-05) | 2.90e-05 ± 2.10e-05 (1.06e-05 - 4.35e-05) |
| Vimentin in μm<sup>2</sup> (IQR) | 7.92e+07 ± 9.68e+07 (1.77e+07 - 8.63e+07) | 1.16e+08 ± 7.77e+07 (7.17e+07 - 1.61e+08) |
| Ki67 positive cells per 1/μm<sup>2</sup> (IQR) | 2.10e-04 ± 2.02e-04 (7.12e-05 ± 3.35e-04) | 5.21e-04 ± 4.49e-04 (1.91e-04 - 8.38e-04) |

### Low and high glioma grades differ in their fluidity properties not in sitffness

Significant differences in tissue microstructure were observed between WHO grade 4 and lower-grade gliomas (grades 2-3). Grade 4 gliomas demonstrated significantly lower *φ* and ADC compared to grade 2 and 3 gliomas (0.48 ± 0.07 rad vs. 0.56 ± 0.1 rad, p < 0.05, and 1073 ± 245 × 10⁻⁶ mm²/s vs. 1522 ± 338 × 10⁻⁶ mm²/s, p < 0.01, respectively). SWS and PR showed no significant differences between groups (1.16 ± 0.12 m/s vs. 1.17 ± 0.25 m/s, and 0.81 ± 0.15 m/s vs. 0.70 ± 0.21 m/s, respectively; both p > 0.05).

### High nuclear aspect ratio in high-grade glioma

AR was significantly higher in grade 4 gliomas compared to grade 2 and 3 gliomas (1.53 ± 0.09 vs. 1.43 ± 0.07, p < 0.01). No significant difference was observed for AR variance (0.15 ± 0.05 vs. 0.11 ± 0.04, p=0.052). Vimentin and reticulin expression showed no significant differences between grade 4 and lower-grade gliomas (116 ± 78 × 10^-6^ µm² vs. 79 ± 97 × 10^-6^ µm², p=0.3, and 2.9 ± 2.0 ×10^-5^ 1/ µm² vs. 1.48 ± 1.0 ×10^-5^ 1/ µm², p = 0.055, respectively). Ki-67 expression was significantly higher in grade 4 gliomas (5.21 ± 4.49 ×10^-4^ 1/ µm² vs. 2.1 ± 2.02 ×10^-4^ 1/ µm², p < 0.05). Box plots of group analyses are presented in Figure 3.

**Figure 3:**
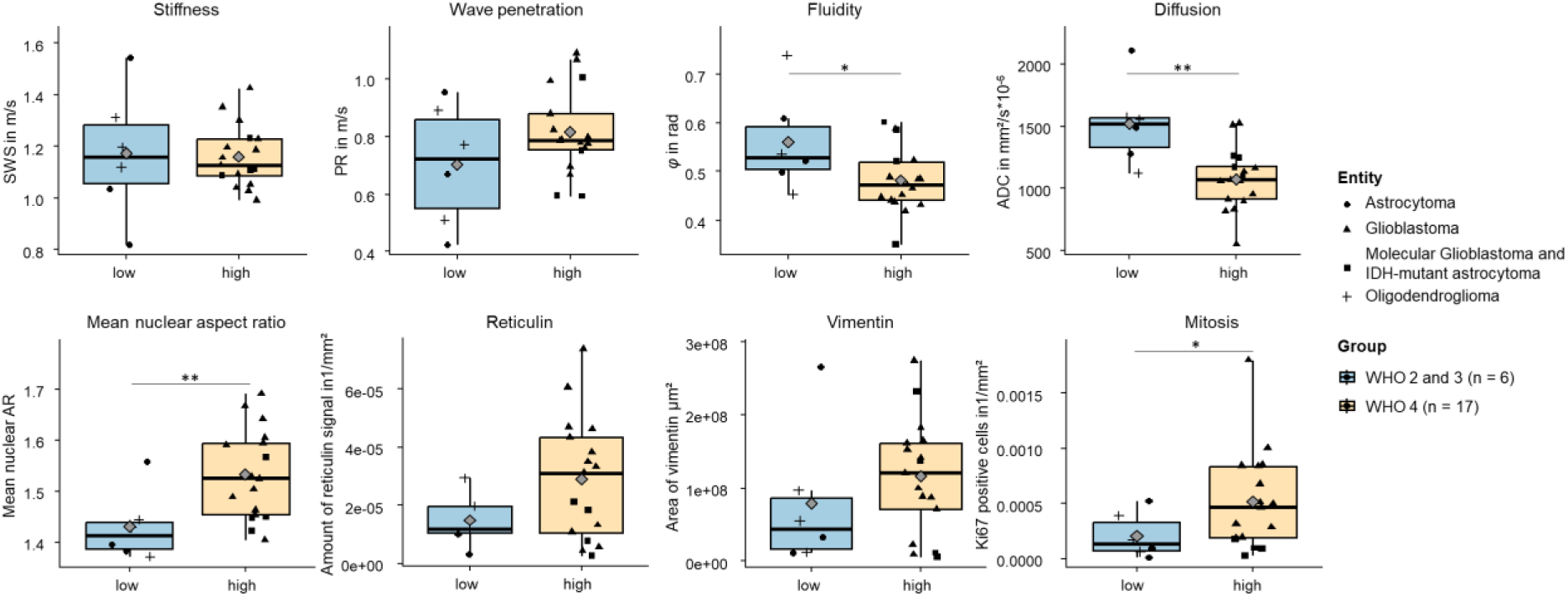
Group statistical plots of mechanical parameters, diffusivity, and quantified histological features comparing WHO grade 4 gliomas with lower-grade gliomas (grades 2–3). Sample sizes are indicated above each box. Mean values are shown as grey diamonds. Statistical significance was assessed using two-sided t-tests or Wilcoxon signed-rank tests, with corresponding p-values reported. *p<0.05; **p<0.01.

### Enhanced histological vimentin levels are associated with reduced MRE viscosity

Considerable variability in mechanical and histological parameters was observed within each glioma grade group, prompting further investigation into the correlations between tissue microstructure and histological characteristics. *φ* demonstrated significant correlations with both AR mean and variance (r = −0.63, p < 0.01; r = −0.47, p < 0.05, respectively), while PR was positively correlated with mean AR (r = 0.44, p < 0.05) and ADC was negatively correlated with mean AR (r = −0.45, p < 0.05). SWS and PR exhibited positive correlations with vimentin area (r = 0.57, p < 0.01; r = 0.75, p < 0.001, respectively), whereas *φ* showed a significant inverse correlation with vimentin expression (r = −0.56, p <0.001). Furthermore, PR and *φ* were correlated with reticulin expression (r = 0.53, p < 0.05 and r = -0.44, p < 0.05, respectively) indicating a better wave penetration into areas of reticulin collagen fiber density. Ki-67-labelled mitotic activity was negatively associated with ADC (r = −0.50, p < 0.05) consistent with the notion of reduced water diffusivity in cell-rich tumor areas(*39, 40*). Ki-67 showed a weak negative correlation with *φ* (r = -0.47, p < 0.05). Corresponding scatter plots depicting these relationships are provided in Figure 4. Correlation analysis revealed no significant associations between SWS and AR, reticulin, or Ki-67, or between PR and Ki-67 (all p > 0.05).

**Figure 4:**
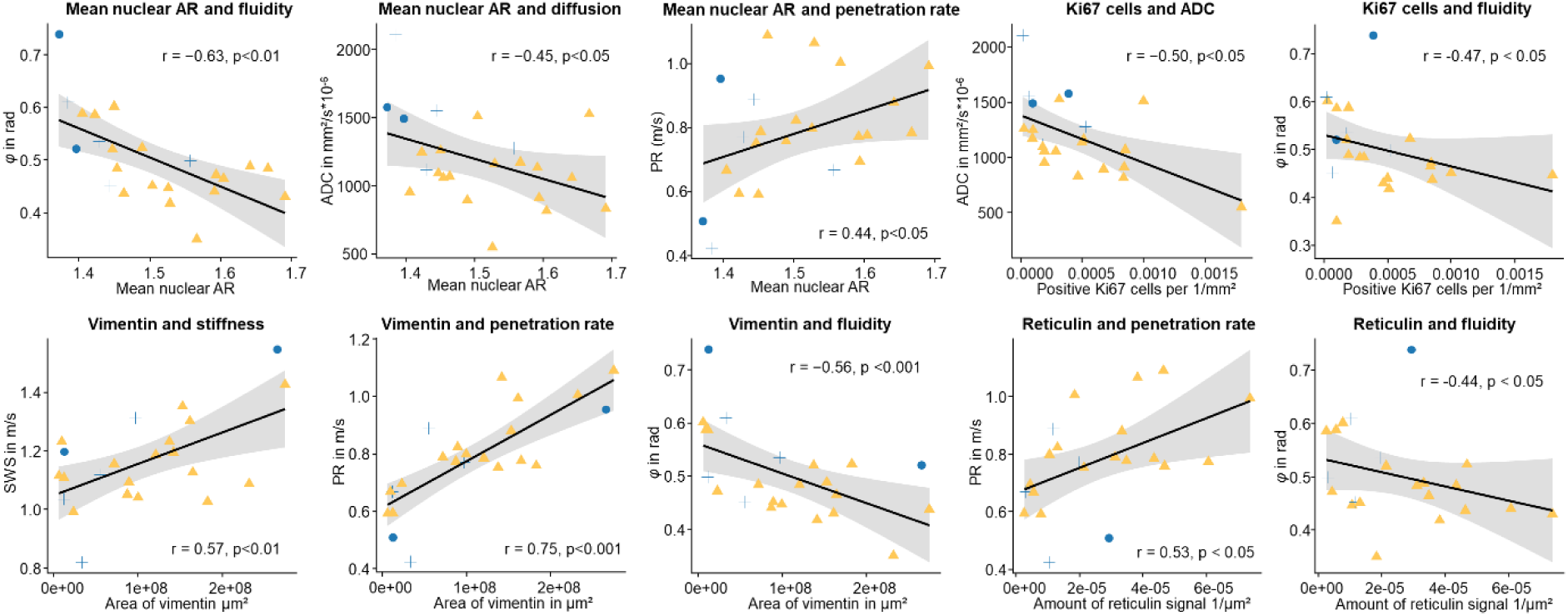
Correlation analyses between histological markers and MRI-MRE parameters. Shaded regions represent 95% confidence intervals from linear regression. Data points are colored by WHO grade (blue for grades 2–3, yellow for grade 4) and shaped according to grade classification (grade 2, grade 3, grade 4). Reported r- and p-values were derived based on linear models.

### Vimentin-to-MRE viscosity correlations are retained in WHO-4 glioma

To account for the unique molecular and biological characteristics of WHO grade 4 gliomas, a subgroup correlation analysis was performed focused on only this group (n = 17, supplemental figure S1). In this subset, *φ* retained significant negative correlations with mean AR and vimentin area (r = –0.55, p < 0.05; r = –0.67, p < 0.01, respectively), whereas PR remained positively correlated with vimentin area and reticulin expression (r = 0.72, p < 0.01; r = 0.52, p < 0.05, respectively).

## Discussion

Combining novel cerebral MRE-based tomoelastography with quantitative histology provided evidence for the key role of vimentin expression in tissue-level biomechanical profiles of in-vivo gliomas. Among all histopathological markers tested in this study, vimentin expression and AR showed the most robust association with MRE parameters. The unique viscoelastic influence of vimentin in polymer solutions with actin demonstrated a reduced fluidity (decreasing loss angle *φ*) and viscosity (increasing PR) over increasing vimentin concentration and provided a mechanistic explanation for the typical soft-solid MRE signature of glioma in patients.(*5, 21, 41*)

In addition and consistent with previous knowledge of vimentin-promoted cell motility from microscopy and single cell tracking, we observed a correlation between vimentin expression and nucleus aspect ratio, which has recently been proposed as a marker of cancer cell motility.(*27*) The combined sensitivities of MRE stiffness, fluidity and viscosity to vimentin expression and MRE fluidity to nucleus shape could provide information on in-vivo tissue biomechanical changes intrinsically linked to proliferation and spread of aggressive cancer cells in glioma.

Unlike many extracranial tumors, in-vivo gliomas are often relatively soft and arise within the already soft background of normal brain tissue, rendering stiffness alone rather insensitive for the detection of human brain tumors.(*42*) This unique soft signature of brain malignancies has been detected by clinical and preclinical MRE independent of the dynamic ranges probed (*41, 43–45*). While clinical MRE is typically performed in a range between 20 to 60 Hz, preclinical MRE in rodent tumor models operates in the kHz-range.(*15*) Consequently, clinical MRE has shown sensitivity to dynamic interactions related to fluid transport, slip or mechanical friction, primarily based on viscosity and fluidity-related parameters, whereas mouse MRE is more dominated by the intrinsic rigidity of mechanical networks and number of cross-links that determine stiffness.(*15*) This sensitivity gap may explain why MRE in tumor mouse models report stiffness as a key parameter that increases with cell and microvessel density,(*44*) cell infiltration,(*46*) while stiffness decreases due to ECM remodeling(*47, 48*) or degraded myelin structures.(*49*)

In contrast, many clinical MRE studies in human brain tumors have demonstrated the importance of viscosity-related parameters such as loss modulus,(*21*) damping ratio(*50*) or fluidity.(*5, 17, 18*) Sauer et al. investigated a variety of brain tumors using combined in-vivo clinical MRE and ex vivo optical stretcher (OS) measurements of suspended cells obtained after tumor resection.(*51*) This one-to-one comparison revealed that in vivo bulk viscosity, rather than stiffness, predicts cellular stiffness and cytoskeletal deformability.(*51*) Specifically, tumor tissue with low bulk viscosity contained softer cells, whereas more viscous tumor tissue harbored relatively stiff cells. These observations suggest that cancer cells adapt their mechanical behavior to the surrounding microenvironment in vivo and that this mechanical profile is preserved in isolated suspended cells.(*8*) It has been proposed that such mechano-sensitivity enables cancer cells to overcome densely packed, jammed conditions by collective fluidization through unjamming transitions.(*13*) Remarkably, unjamming transitions are associated with shape changes towards more elongated cell geometries, which are also reflected in an increased AR.(*26*) The observed negative correlation between AR and *φ* in this study is consistent with previous reports in ex vivo tumor models of human-derived prostate cancer cell lines.(*52*) There, it was discussed that differences in cell size between prostate tumors lead to cell-density-driven jamming, which can mask cell-shape related tissue fluidization. This interpretation was supported by the fact that ADC was reduced in samples with high AR indicating densely clustered cells, with, however, shape-induced motility. Also in our study, ADC was reduced with AR. Furthermore, tissue fluidization due to cancer cell unjamming appears to be counterbalanced by vimentin expression, which, as shown by the phantom experiments, results in lower fluidity values.

In dense tumor niches, cancer cells are routinely exposed to mechanical stress from growth-induced solid stress and from fluid pressure due to aberrant, leaky vessels formed by tumor neoangiogenesis.(*53*) To survive and proliferate, cancer cells need to resist mechanical stress by overexpressing vimentin.(*30*) Vimentin acts as a ‘safety belt’ protecting against compressive stress and preserves mechanical integrity by enhancing cell elastic behavior.(*33*) Hu et al. demonstrated that the stretchability and hyperelastic nature of the vimentin intermediate filament networks allows cells resist more than 300% strain by effectively transmitting it from a stress concentrated area to a larger zone in the cell.(*54*) Beyond cytoskeletal force propagation, vimentin can exhibit ECM stiffness-dependent effects on other cytoskeletal components (i.e., the actomyosin and microtubule networks) and therefore directly modulate traction forces and control cell motility. For example, wild type cells of fibroblasts containing vimentin spread farther than vimentin depleted cells when exposed to soft substrates such as brain ECM.(*28*)

Several studies have correlated MRE-derived viscoelastic parameters in glioma and glioblastoma with histopathological markers. In orthotopic brain tumor models, Jamin et al. showed that both elasticity and viscosity correlate positively with tumor cell density and microvessel density indicating that stiffness and viscosity primarily reflect cellularity and vascular proliferation in these glioma-like lesions.(*44*) Li et al. demonstrated strong correlations between picrosirius-red staining and both elasticity and viscosity suggesting that collagen-rich stroma is a key determinant of the tumor biomechanical phenotype, including glioblastoma xenografts.(*55*) In GBM mouse models, Schregel et al. reported that softer GBM regions in MRE correspond to necrosis and sparsely viable tumor, while relatively stiffer regions match densely packed tumor cells and blood vessels in histology.(*45*) Similarly, Janas et al. found that tumors were softer than contralateral brain regions and reduced φ values at later stages correlated with marked vascularization and elevated glycosaminoglycan content.(*47*) In human glioblastoma, Svensson et al. combined MRE with stereotactic biopsies and found that locally increased stiffness co-localizes with ECM reorganization and ECM/adhesion-related gene-expression associated with shorter survival.(*56*) Despite these insights into glioma microarchitecture and macroscopic MRE properties, none of these studies established a mechanistic link between tissue mechanics and the aggressiveness or invasive behavior of cancer cells. By focusing on vimentin expression, nuclear elongation, and novel MRE markers such as penetration rate, our study is, to our knowledge, the first to demonstrate this link in vivo and, at the same time, to offer a new window into tumor biology through high-resolution tomoelastography.

Our study has limitations. First, tomoelastography provides macroscopic, voxel-averaged measures of stiffness, viscosity and fluidity, whereas histology and nuclear shape analysis are based on small, spatially restricted biopsy samples, introducing potential sampling and co-registration bias. Second, the in vitro actin-vimentin phantoms recapitulate key cytoskeletal features but cannot fully mimic the complexity of the glioma microenvironment, including heterogeneous ECM composition, vasculature and immune infiltrates. Finally, the MRE sequence and specific reconstruction parameters may be further optimized to achieve even higher spatial resolutions as recently proposed by Herthum et al.(*57*).

In summary, we combined multifrequency cerebral tomoelastography with quantitative histopathology and viscoelastic phantoms to reveal that in vivo glioma mechanics are tightly linked to vimentin-driven cytoskeletal remodeling and nuclear elongation. Decreased tumor fluidity and viscosity emerge as soft-solid hallmarks of glioma that scale with vimentin expression, nuclear aspect ratio and, by inference, collective cancer cell migration. By mechanistically linking macroscopic viscoelastic signatures to microscopic cytoskeletal architecture, our work bridges the gap between radiological imaging and multiscale mechanics in human brain tumors. These findings suggest high-resolution tomoelastography as a non-invasive biomechanical window into glioma aggressiveness and may open new avenues for stratifying patients and monitoring therapies based on the physical state of their tumors.

## Methods

### Phantoms

To study the fundamental viscoelastic behavior of vimentin mixed with other cytoskeletal proteins, we evaluated data previously reported by Golde et al.(*33*). Phantom preparation and rheometric measurements are briefly summarized in the following.

#### Preparation of actin/vimentin composite phantoms

G-actin was purified from rabbit skeletal muscle and stored at −80 °C in G-buffer (2 mM sodium phosphate, pH 7.5, 0.2 mM ATP, 0.1 mM CaCl₂, 1 mM DTT, 0.01% NaN₃) until use. Human vimentin was recombinantly expressed in E. coli, purified from inclusion bodies and refolded by stepwise dialysis from 8 M urea into 2 mM sodium phosphate buffer (pH 7.5). Immediately before experiments, monomeric actin and vimentin were mixed on ice at the desired concentrations in low-ionic-strength buffer. Polymerization and network formation were initiated by adding 1/10 volume of 10× F-buffer (20 mM sodium phosphate, pH 7.5, 1 M KCl, 10 mM MgCl₂, 2 mM ATP, 10 mM DTT), yielding fully co-polymerized actin/vimentin gels with interpenetrating filament networks.

#### Rheometric measurement of complex shear modulus

The viscoelastic properties of the composite phantoms were measured using a strain-controlled ARES rheometer (TA Instruments) equipped with a 40 mm plate–plate geometry and a gap of 140 µm. Samples were mixed on ice and immediately loaded between the plates; polymerization proceeded in situ for 2 h at 25 °C. To minimize edge and evaporation artifacts, F-buffer was applied around the plate rim and the chamber was sealed with a cap containing moist sponges. Network assembly was monitored by small-amplitude oscillatory shear (time sweep, 1 Hz, 5% strain, one measurement per 60 s) until the modulus reached a steady state. The linear viscoelastic moduli were then obtained from frequency sweeps (0.01–10 Hz, 5% strain, 20 frequencies per decade), yielding the storage modulus G′ and loss modulus G″ from the complex shear modulus G*(ω).

### Patients

Twenty-four consecutive patients with suspected glioma and no history of prior surgery, radiotherapy, or chemotherapy were prospectively enrolled. One patient was excluded after histopathology showed no tumor, leaving 23 patients for analysis. Hereof 17 tumors were WHO grade 4 and were 6 WHO grade 2–3 gliomas. All patients underwent preoperative MRI including multifrequency MRE. The study protocol is in accordance with the Declaration of Helsinki and was approved by the local ethics committee (EA1/248/19), and all patients provided written informed consent.

#### MRI and MRE acquisition

Imaging was performed on a 3-Tesla MRI system (Magnetom Lumina, Siemens Healthineers, Erlangen, Germany) using a 32-channel head coil. Conventional anatomical imaging included a 3D T1-weighted MPRAGE sequence for structural reference as well as a 2D FLAIR sequence. Water diffusion was quantified by means of apparent diffusion coefficient (ADC) using a spin-echo echo-planar imaging sequence with 0 and 1000 b-values. Brain MRE was acquired with a single-shot spin-echo EPI sequence as previously described for cerebral tomoelastography.(*36*) Scan parameters were: repetition time 4,700 ms, echo time 70 ms, field of view 202 × 202 mm², acquisition matrix 126 × 126, voxel size 1.6 × 1.6 × 2.0 mm³, and 40 contiguous axial slices. Motion-encoding gradients with an amplitude of 34 mT/m and a duration of 28 ms were applied along three orthogonal directions and synchronized to the mechanical vibration. Shear waves were induced by a pneumatic driver system consisting of two cushions coupled to a rigid transmission plate positioned underneath the patient’s head within the head coil, and operated in opposed-phase mode to minimize compression waves. Multifrequency vibrations at 20, 25, 30, and 40 Hz were applied. For each frequency, eight equally spaced time steps over one vibration period were sampled, yielding a multifrequency, three-directional displacement dataset per patient. Total acquisition time for the 3D multifrequency MRE dataset (40 slices, 4 frequencies, 8 time steps, 3 motion-encoding directions) was approximately 8 minutes. The MRE setup along with wave images is shown in Figure 5.

**Figure 5.**
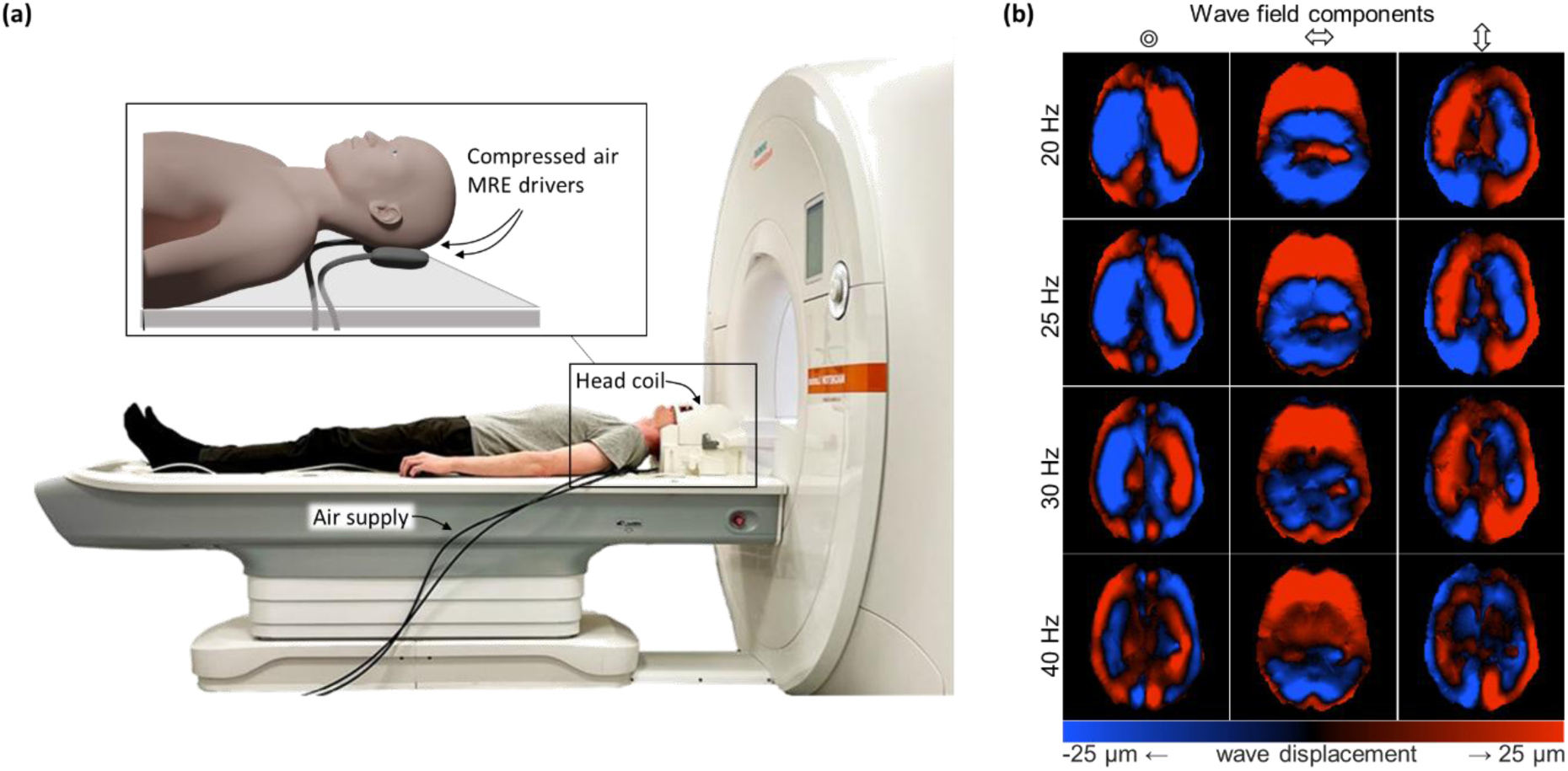
Experimental setup and brain MRE wave fields. **(a)** Clinical 3T MRI setup with a 32-channel head coil and two compressed-air MRE drivers placed beneath the patient’s head and connected to an external air supply. The inset illustrates the positioning of the head drivers relative to the occiput. **(b)** Representative MRE displacement fields in three orthogonal motion-encoding directions at driving frequencies of 20, 25, 30, and 40 Hz. Wave amplitudes are color-coded from −25 µm (blue) to +25 µm (red).

#### Tomoelastography reconstruction

Before MRE processing, motion artifacts were corrected by retrospective image registration using the open-source elastix toolbox.(*58*) For each subject, the magnitude image of the first time frame was used as a reference, and all remaining time frames, motion-encoding directions and drive frequencies were registered slice-wise to this reference volume using a standard multi-resolution affine/B-spline scheme as described by Shahryari et al.(*59, 60*). The resulting deformation fields were then applied to the corresponding complex wave images to obtain motion-corrected displacement data for tomoelastography inversion. The tomoelastography pipeline used based on wavenumber-based multifrequency dual elasto-visco (k-MDEV) inversion described in detail in Herthum et al.(*36*) and publicly available under https://bioqic-apps.charite.de.(60) In brief, after temporal Fourier transformation, complex displacement fields were decomposed into eight directional shear wave components and filtered using a bandpass filter to suppress longitudinal waves and noise. For each frequency, local complex wavenumbers were estimated from the spatial phase gradients of the complex wave images in each slice. Frequency-specific shear wave speed (SWS) and penetration rate (PR) maps were then obtained and combined across drive frequencies using amplitude-weighted averaging to increase robustness. SWS maps (in m/s) were interpreted as surrogate measures of tissue stiffness, while PR (m/s) reflected viscous damping. Phase angle *φ* (rad) of the complex modulus was reconstructed from SWS and PR by

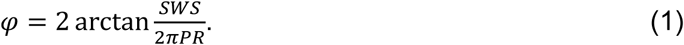

As suggested in Streitberger et al.(*5*), *φ* was interpreted as tissue fluidity as it increases from 0 for pure elastic solids to *π*/2 for viscous fluids which do not support shear forces. No further model-based conversion to storage or loss moduli was performed. Contrast enhanced MPRAGE, FLAIR and apparent diffusion coefficient (ADC) from diffusion weighted images (DWI) were co-registered to MRE data using ITKsnap for region-of-interest analysis of the tumors.(*61*)

### Histological stains and quantification

Formalin-fixed, paraffin-embedded tumor samples were sectioned and stained using standard protocols for hematoxylin-eosin stain, reticulin silver stain and immunohistochemical stains. The following primary antibodies were used: Ki-67 antigen (DAKO M 7240, clone Mib-1, 1:100) and vimentin antigen (DAKO M 7020, clone Vim3B4, 1:400). Whole-slide images were processed using a custom Python script based on the OpenSlide library. Each slide was divided into non-overlapping tiles of 5000 × 5000 pixels at full resolution, and tiles were screened for background by thresholding the mean RGB intensity; tiles with more than 50% background pixels were discarded. The remaining tissue-containing tiles were saved as TIFF files and organized into directories by stain and case identifier. Hematoxylin-eosin staining was performed for general morphology and cell nucleus shape analysis. Nuclear aspect ratio (AR) was quantified based on digitized histological slides using the CeNuS (cell and nucleus shape) framework as described by Gottheil et al.(*26*). Briefly, nuclear contours were automatically segmented by a deep learning-based instance segmentation algorithm, and for each detected nucleus an equivalent ellipse was fitted to extract the major and minor axes. The nuclear AR was defined as the ratio of the major to the minor axis, yielding a dimensionless measure of nuclear elongation; for each region of interest (ROI), the distribution of AR values was computed and summarized (e.g., by mean AR) for subsequent correlation with imaging and clinical parameters.(*62*) Vimentin expression, reticulin fiber content, and Ki-67 proliferation index were quantified by digital image analysis, yielding mean labeling indices or area fractions for each marker. In brief, we perform for each slide a custom color-deconvolution based on the algorithm proposed by Macenko et al.(*63*), yielding the spatially resolved positive staining intensities (typically brownish) and the negatively stained structures (typically blueish). Representative histological images along with quantified markers and region of interests (ROI) are shown in Figure 6.

**Figure 6.**
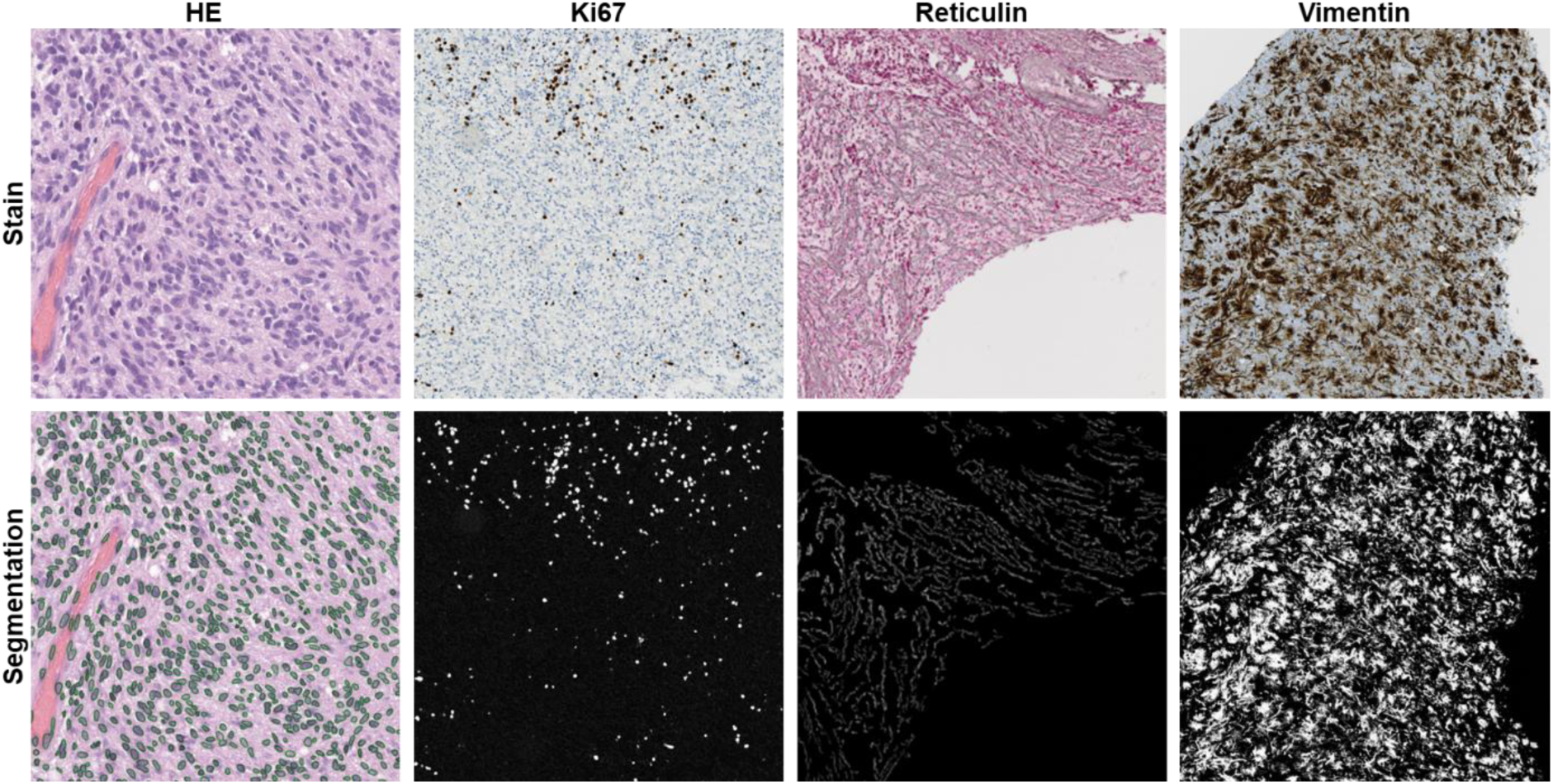
Histological stains and quantitative image analysis. Representative examples of hematoxylin-eosin (H&E), Ki-67, reticulin, and vimentin stains (top row) and corresponding segmentations used for quantitative analysis (bottom row). For H&E, individual nuclei were automatically segmented (green overlays) and nuclear shapes were characterized by their aspect ratio (AR) to quantify nuclear elongation. For Ki-67, reticulin, and vimentin, binarized masks illustrate the segmented positive signal used to compute labeling indices or area fractions for each marker.

### Statistical analysis

The region of interest (ROI) was delineated on the preoperative anatomical MRI (contrast enhanced T1-weighted MPRAGE and FLAIR) based on the region identified by the neurosurgeon after tumor resection as corresponding to the tissue submitted for pathological examination. The ROIs were then co-registered and transferred to the corresponding tomoelastography maps, ensuring spatial alignment between morphological and mechanical data. Mean values of SWS, PR and *φ* were extracted from each ROI for subsequent statistical analysis. Statistical analysis was performed using non-parametric and parametric tests as appropriate. WHO grade 2 and 3 gliomas were grouped as “low”, whereas WHO grade 4 gliomas were grouped as “high”. Group differences were assessed with Kruskal–Wallis tests and, where applicable, two-sided t-tests or Wilcoxon signed-rank tests. Associations between mechanical parameters and clinical or imaging variables were evaluated using Pearson and Spearman correlation coefficients. Normality of residuals was checked by visual inspection of quantile–quantile plots.

## Data Availability

MRE data produced in the present study are available upon reasonable request to the authors

## Acknowledgments

Mehrgan Shahryari is participant in the BIH Charité Junior Digital Clinician Scientist Program funded by the Charité - Universitätsmedizin Berlin, and the Berlin Institute of Health at Charité (BIH). This study was supported by the German Research Foundation (DFG, FOR5628 grant No. 513752256, CRC1340, GRK2260 BIOQIC, CRC1540 grant No. 460333672).

---

**Supplemental figure S1.**
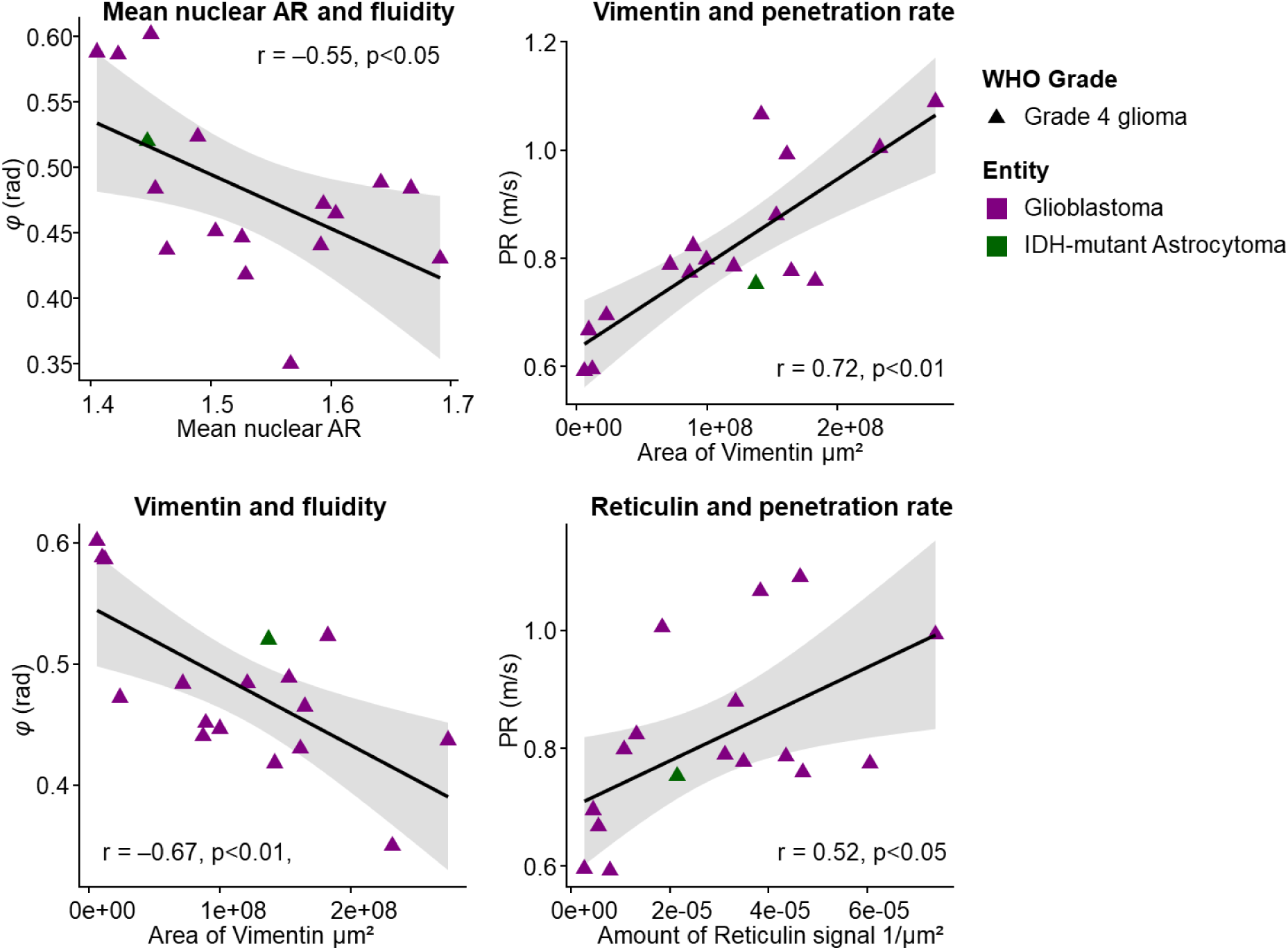
Subgroup correlation analyses within WHO grade 4 gliomas. Shaded regions represent 95% confidence intervals from linear regression. Data points are colored uniformly for grade 4 and shaped to indicate individual cases. Reported r- and p-values derive from these models.

